# Interpretable Artificial Intelligence for COVID-19 Diagnosis from Chest CT Reveals Specificity of Ground-Glass Opacities

**DOI:** 10.1101/2020.05.16.20103408

**Authors:** Anmol Warman, Pranav I. Warman, Ayushman Sharma, Puja Parikh, Roshan Warman, Narayan Viswanadhan, Lu Chen, Subhra Mohapatra, Shyam S Mohapatra, Guillermo Sapiro

## Abstract

1

**Background:** The use of CT imaging enhanced by artificial intelligence to effectively diagnose COVID-19, instead of or in addition to reverse transcription-polymerase chain reaction (RT-PCR), can improve widespread COVID-19 detection and resource allocation.

**Methods:** 904 axial lung window CT slices from 338 patients in 17 countries were collected and labeled. The data included 606 images from COVID-19 positive patients (confirmed via RT-PCR), 224 images of a variety of other pulmonary diseases including viral pneumonias, and 74 images of normal patients. We developed, trained, validated, and tested an object detection model which detects features in three categories: ground-glass opacities (GGOs) for COVID-19, GGOs for non-COVID-19 diseases, and features that are inconsistent with a COVID-19 diagnosis. These collected features are passed into an interpretable decision tree model to make a suggested diagnosis.

**Results:** On an independent test of 219 images from COVID-19 positive, a variety of pneumonia, and healthy patients, the model predicted COVID-19 diagnoses with an accuracy of 96.80 % (95% confidence interval [CI], 96.75 to 96.86), AUC-ROC of 0.9664 (95% CI, 0.9659 to 0.9671), sensitivity of 98.33% (95% CI, 98.29 to 98.40), precision of 95.93% (95% CI, 95.83 to 95.99), and specificity of 94.95% (95% CI, 94.84 to 95.05). On an independent test of 34 images from asymptomatic COVID-19 positive patients, our model achieved an accuracy of 97.06% (95% CI, 96.81 to 97.06) and a sensitivity of 96.97% (95% CI, 96.71 to 96.97). Similarly high performance was also obtained for out-of-sample countries, and no significant performance difference was obtained between genders.

**Conclusion:** We present an interpretable artificial intelligence CT analysis tool to diagnose COVID-19 in both symptomatic and asymptomatic patients. Further, our model is able to differentiate COVID-19 GGOs from similar pathologies suggesting that GGOs can be disease-specific.

## 2 Introduction

Since the onset of the COVID-19 pandemic, over 4,000,000 cases have been confirmed and over 280,000 patients have died. Currently, patients are diagnosed by Reverse Transcription Polymerase Chain Reaction (RT-PCR) from nasopharyngeal or throat swab.^1,2^ However, widespread testing is still not readily accessible despite the acceptance that it is critical to rapidly and accurately test individuals to control a pandemic.^3–5^ Further, a major disadvantage of current tests include lack of sensitivity, which results in a 10–30% false negative rate.^6–8^ The shortage of adequate testing has pushed researchers to study complementary options including antibody testing and computed tomography (CT).^9–11^ Notably, recent studies have shown that CT can be used to diagnose COVID-19 and that this can be more sensitive than RT-PCR testing.^10,11^ CT as a testing platform is further beneficial as chemical contaminations which caused shortages of RT-PCR testing are not problems and there is an existing infrastructure for CT in almost every hospital. However, using CT for active screening and diagnosis is currently complicated by the absence of guidelines for detecting COVID-19, which can appear identical to many pulmonary conditions.^6,12–14^

CT imaging combined with an artificial intelligence (AI) system capable of not only diagnosing^*^ COVID-19 but also doing so with few false positives has the potential to improve clinical response and patient outcome. Recent publications have proposed AI classifiers for COVID-19 diagnosis; however, these tools provide little understanding of how decisions were made, which makes confirming the diagnosis as challenging of a process as diagnosing a patient in the first place. Further, these tools are limited in their ability to differentiate COVID-19 from phenotypically similar diseases.^15–18^

Central to the diagnostic process are ground-glass opacities (GGOs), the radiological finding that indicates the presence of not only COVID-19 but also many similar diseases.^6,12–14^ While it is conventionally understood that GGOs are non-specific, recent literature suggests that there may be subtle variations of GGO manifestation between broad disease classes.^19^ We hypothesized that an AI system may be sensitive to these nuances and capable of distinguishing COVID-19 GGOs from GGOs of other diseases in the non-opportunistic infection class. To preserve the integrity of clinical decision making, we propose that such a system should also offer transparency into the usual “black-box” AI tools.

Here, we report an AI diagnostic system that is able to identify, from CT scans, COVID-19 in a highly accurate, sensitive, and precise manner. The system is able to locate and label GGOs in a CT slice as related or unrelated to COVID-19, and it recognizes features inconsistent with a COVID-19 diagnosis, namely lobar consolidations, cavitations, and pleural effusions.^12^ The system combines all labeled features in an interpretable decision tree to suggest a diagnosis for radiological review and supports its proposal by presenting visuals with radiological evidence highlighted. The results reveal the feasibility of COVID-19 diagnosis regardless of symptomatic presence and demonstrates the heterogeneity of GGOs within non-opportunistic infections.

## 3 Materials and Methods

### 3.1 Study Design

The aim of the study was to develop and evaluate the performance of an artificial intelligence system for COVID-19 diagnostic purposes. The model was trained, validated, and tested on CT scans of 2,116 COVID-19 GGO instances (labeled COVID-19), 569 instances of non-COVID-19 GGOs (labeled non-COVID GGO), and 143 instances of features which are inconsistent with a COVID-19 diagnosis (labeled DQ). These were identified across 606 COVID-19 positive and 224 COVID-19 negative images. All COVID-19 positive patients were reported as confirmed by RT-PCR testing. Normal chest CT scans were collected from 74 patients for false positive control. All data was obtained, with permission, from public domain sources and therefore IRB was not deemed necessary for the study (Table S1).

To offer transparency, an object detection architecture was used so bounding boxes around detected features could be generated for radiological review. For training data, each important region was labeled by two upper-level radiology residents. Subsequently, labeling was confirmed with a board-certified diagnostic radiologist with 10 years of experience to establish consensus. Unless specified otherwise, the data was split in a 7:1:2 ratio between training, validation, and test sets. The test set was refined to only include patients completely independent of the training and validation set. This was done to avoid any unknown correlation between test and train sets. Two countries were also entirely kept in the test set to control for hospital-specific protocols.

### 3.2 Image Acquisition

Retrospectively collected COVID-19 images were obtained from a variety of CT scanners; the most commonly reported scanner type was 64-slice. Only axial lung window images were included for both training and testing groups. The de-identified images were obtained from open-source full CT stacks and materials used for COVID-19 education (Table S1). CT images containing pertinent radiological findings were manually curated from the full stacks by radiologists in order to generate the instance counts described above.

### 3.3 Study Patients

The study included patients with COVID-19, many phenotypically similar pulmonary conditions, and healthy patients representing 17 countries (Table S1). The patients’ CT scans were collected across various hospitals and the de-identified CT stack was uploaded for open-source use. Only patients whose diagnoses were marked “Diagnosis certain” were included in this study to ensure the model was robust.

### 3.4 System Architecture

The YOLOv3 architecture,^20^ currently considered the state-of-art for object detection, was adapted to work as a three-class single-shot detector. Transfer learning has been shown to reduce error and time to convergence;^21^ therefore, the model weights were initialized from a YOLOv3 model that was trained on the COCO dataset^22^ until it achieved a mean average precision (mAP) of 0.579 for a 0.5 Intersection over Union (IoU).

Additionally, a simple decision tree was designed to predict COVID-19 diagnosis based on the presence or absence of the three classes (features) measured by the YOLOv3 model (Fig. S2).

### 3.5 Training

The YOLOv3 model was trained on the aforedescribed train set for 5,000 epochs. Every 1,000 epochs the model was evaluated on the validation set to check for overfitting and to check if an earlier stopping point out-performed the final model (Figs. S1a-f). The best model was chosen and then evaluated in terms of accuracy, sensitivity, precision, specificity, false positive rate, and the Area under the Curve of the Receiver Operating Characteristic (AUC-ROC) for both overall performance on diagnosing COVID-19 and for performance of diagnosing COVID-19 on the test set of data for specific diseases. (For information on calculation methodologies, see Section S1)

### 3.6 Statistical Analysis

A Receiver Operating Characteristic (ROC) curve was calculated using sensitivity and specificity values from each evaluated model instance. By under-sampling the ROC space with only one point, we are underestimating the area under the curve (AUC) for the ROC.^23^ Therefore, the values presented, while strong, represent a lower bound of our AUC-ROC. Other performance metrics complement this.

N-out-of-n bootstrap with replacement was used with images as the resampling unit. For each of the 2000 bootstrap samples, we calculated performance metrics and used this sampling to estimate a 95% confidence interval (CI) for each metric.^24^

## 4 Results

### 4.1 Object Detection Performance

Our object (region) detection system is able to process, detect, and label each image for instances of COVID-19 GGOs, non-COVID GGOs, and features that would disqualify a COVID-19 diagnosis. Run time for detection is less than 20 milliseconds on a Tesla P100-PCIE-16GB GPU, and the system does this accurately with a 0.59 mean average precision (mAP) for a 0.5 threshold for intersection over union.

### 4.2 Performance on External Test Set

The deep learning system and decision tree are combined to make an interpretable classifier.

After validating the model on a set of 112 images, the system was evaluated on the test set. The test data set was representative of the overall data (Table S2). For reported patients, the test set had a 1:1 male to female ratio whose median age was 54.76 (range, 22 to 88).

The performance of the proposed system on the entire dataset was measured with an accuracy of 96.80 % (95% confidence interval [CI], 96.75 to 96.86), AUC-ROC of 0.9664 (95% CI, 0.9659 to 0.9671), sensitivity of 98.33% (95% CI, 98.29 to 98.40), precision of 95.93% (95% CI, 95.83 to 95.99), and specificity of 94.95% (95% CI, 94.84 to 95.05). On a subset of countries and hospitals not represented at all in the training data, our model reported an accuracy of 98.36% (95% CI, 98.29 to 98.43), AUC-ROC of 0.9914 (95% CI, 0.9909 to 0.9917), sensitivity of 98.28% (95% CI, 98.19 to 98.34), precision of 100% (95% CI, 100 to 100), and specificity of 100% (95% CI, 100 to 100). The model was found to perform slightly better on female patients (accuracy: 98.18% [95% CI, 98.13 to 98.29], AUCROC: 0.9500 [95% CI, 0.9484 to 0.9527], sensitivity: 100% [95% CI, 100 to 100], precision: 97.83% [95% CI, 97.77 to 97.96], specificity: 90.00% [95% CI, 89.67 to 90.53]) when compared to the set of male patients (accuracy: 93.85% [95% CI, 93.79 to 94.04], AUC-ROC: 0.9411 [95% CI, 0.9403 to 0.9428], sensitivity: 96.55% [95% CI, 96.27 to 96.58], precision: 90.32% [95% CI, 90.33 to 90.79], specificity: 91.67% [95% CI, 91.69 to 92.08]) (Table 1, Fig. 1). Model performance was also evaluated per subclass of disease type to better understand model biases and it was determined that no subclass had an outlier performance. In all cases, model prediction returned the diagnosis with images that include drawn boxes showing the location of the features the model used to generate its diagnosis (Figs. 2, S4a, S4b). This is a clear example of the interpretability of the proposed model and its value as an aid in diagnosis.

**Table 1:** Performance of the object detection model in conjunction with the decision tree on the validation set, external test set, set of independent countries, and set of asymptomatic patients.

|  | Accuracy<br>(95% CI) | Sensitivity<br>(95% CI) | Precision<br>(95% CI)<br>*percent* | Specificity<br>(95% CI) | False Positive Rate<br>(95% CI) | AUC-ROC<br>(95% CI)<br>*value* |
| --- | --- | --- | --- | --- | --- | --- |
| **Total External Test** | 96.80<br>(96.75-96.86) | 98.33<br>(98.29-98.40) | 95.93<br>(95.83-95.99) | 94.95<br>(94.84-95.05) | 5.050<br>(4.950-5.159) | 0.9664<br>(0.9659-0.9671) |
| **External Test Subsets** |  |  |  |  |  |  |
| Independent Countries | 98.36<br>(98.29-98.43) | 98.28<br>(98.19-98.34) | 100<br>(100-100) | 100<br>(100-100) | 0<br>(0-0) | 0.9914<br>(0.9909-0.9917) |
| Asymptomatic | 97.06<br>(96.81-97.06) | 96.97<br>(96.71-96.97) | - | - | - | - |
| Pneumonias | 94.74<br>(94.52-94.98) | - | - | 94.74<br>(94.52-94.98) | 5.263<br>(5.019-5.476) | - |
| Male | 93.85<br>(93.79-94.04) | 96.55<br>(96.27-96.58) | 90.32<br>(90.33-90.79) | 91.67<br>(91.69-92.08) | 8.333<br>(7.919-8.312) | 0.9411<br>(0.9403-0.9428) |
| Female | 98.18<br>(98.13-98.29) | 100<br>(100-100) | 97.83<br>(97.77-97.96) | 90.00<br>(89.67-90.53) | 10.00<br>(9.468-10.33) | 0.9500<br>(0.9484-0.9527) |
| **Validation** | 98.21 | 100 | 97.40 | 94.59 | 5.41 | 0.9730 |

**Figure 1:**
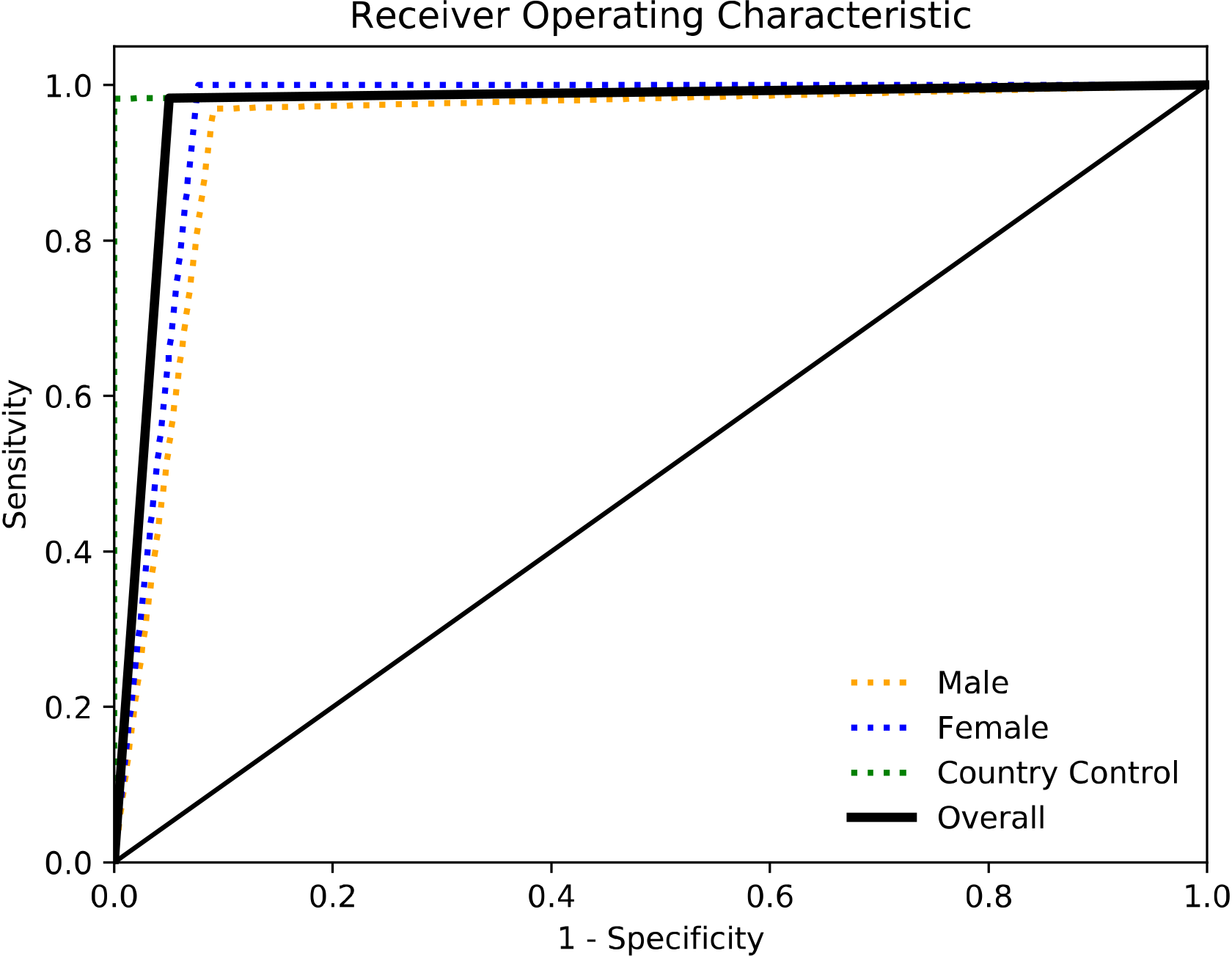
Performance on Test Sets and Controls. The external test set included CT images from 17 countries with diverse pathologies. Our interpretable deep learning system is able to distinguish GGOs of COVID-19 from GGOs of other non-opportunistic infections, with an overall area of the receiver-operating-characteristic curve (AUCs) of 0.9664.

**Figure 2:**
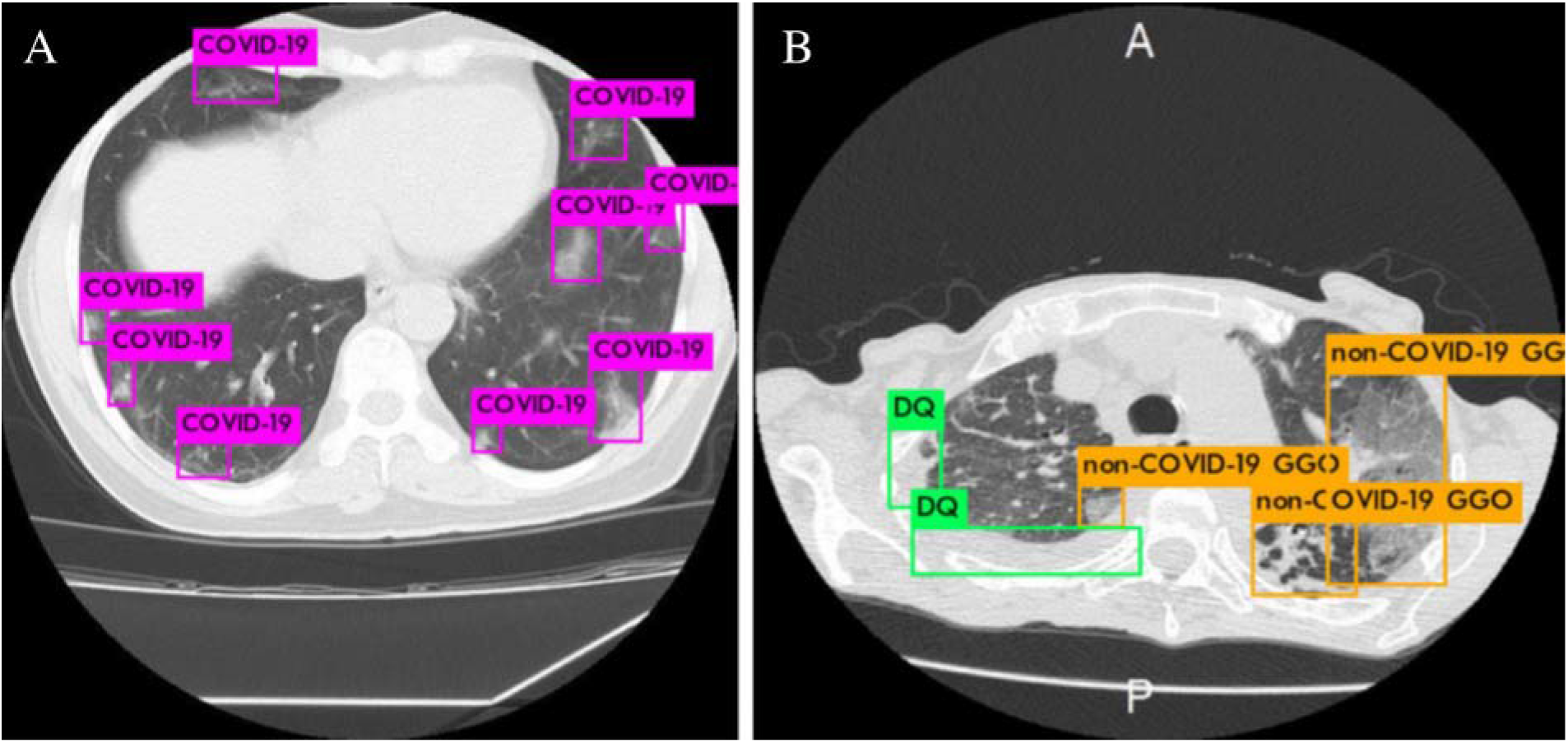
GGO Differentiation and Example Model Prediction. (**A)** A 74-year-old male diagnosed with COVID-19 was scanned. Nine COVID-19 GGOs were labeled and presented along with the suggested diagnosis that the patient was COVID-19 positive. (**B)** A 53-year-old woman diagnosed with lipoid pneumonia and presented symptoms similar to COVID-19. The model was able to detect and differentiate the non-COVID GGO from a COVID radiological finding. Additionally, the model observed the presence of a DQ feature, specifically a cavitation.

### 4.3 Classification of Asymptomatic COVID-19 Patients

In an effort to understand the robustness of our proposed model, we tested it on 34 images from 14 patients (7 male, 7 female, median reported age of 61 ± 7) who were asymptomatic of COVID-19. The model was able to diagnose these 14 patients with an accuracy of 97.06% (95% CI, 96.81 to 97.06) and a sensitivity of 96.97% (95% CI, 96.71 to 96.97) (Table 1). As before, model prediction returned the diagnosis with images that include drawn boxes showing the location of the features the model used to generate its diagnosis (Fig. 3).

**Figure 3:**
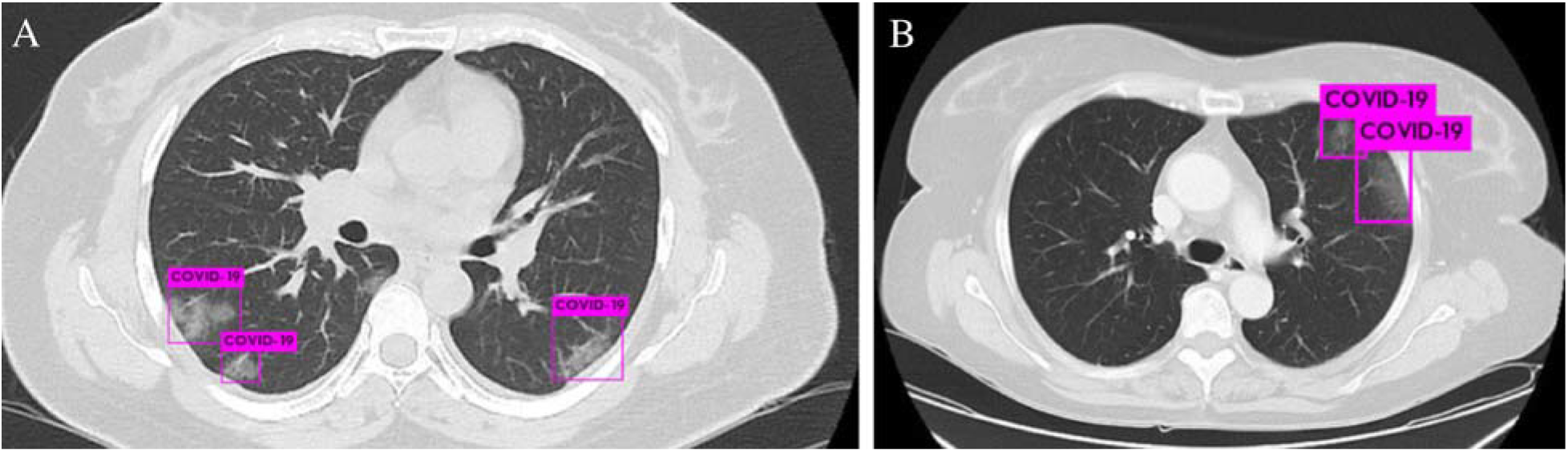
Radiological Findings of COVID-19 in Asymptomatic Patients. (**A)** A 50-year-old female had traveled to Algeria and made contact with a COVID-19 positive patient. Four days later this scan was taken, on which the model is able to indicate COVID-19 GGOs with tight bounding boxes. Confirmed as COVID-19 positive by RT-PCR. (**B)** A 60-year-old female had a routine follow up scan for oncological treatment of GI tract cancer with no known metastases. Although clinically well, the patient underwent RT-PCR testing which confirmed the presence of COVID-19, as the model was able to detect.

### 4.4 Review of Errors

To gain further insight into the model, each erroneous diagnosis was adjudicated, and it was determined that in each case the incorrect diagnosis was atypical. For example, an asymptomatic patient diagnosed as COVID-19 negative, radiologist inspection of the CT slice showed no visible COVID-19 features due to poor photo quality.

In all cases, further review and analysis were able to determine the cause of the error (Figs. S3ac).

## 5 Discussion

In this study, we introduce an interpretable deep learning system for CT analysis that is highly accurate, sensitive, and precise in diagnosing COVID-19. The system combines an object detection model to identify features that are explicitly labeled with an interpretable decision tree to make a COVID-19 diagnosis.

We find that such a system performs equally well on symptomatic and asymptomatic patients and can differentiate between GGOs of COVID-19 and GGOs of alternate pathological conditions, particularly viral pneumonias. On an external test set, our model reported excellent accuracy, AUC-ROC, sensitivity, precision, and specificity. This was also the case when the model was tested on a subset of countries and hospitals not represented in the training data, suggesting generalization over hospital and country-specific imaging practices.

The results demonstrate that the model is able to differentiate COVID-19 GGOs from non-COVID GGOs, which has large radiological and diagnostic ramifications. Specifically, this suggests that there are phenotypical differences between various manifestations of GGOs of non-opportunistic infections and that artificial intelligence models, such as the one presented here, are sensitive enough to distinguish them. In line with this, we found that our model was able to distinguish COVID GGOs from those of diseases not initially trained on, including miliary tuberculosis, asbestos-related pleural disease, and pulmonary metastases related to various primary malignancies.

Our model can diagnose COVID-19 even in asymptomatic patients – cases that are essential to accurately monitor and contain the spread of the virus. Further, using CT scans allow for incidental findings of COVID-19 in these patients, which could lead to the implementation of more effective public quarantine policies.

By making our system interpretable, revisions of model diagnosis and model errors become a simple process. Our results indicate that the model was capable of generating tight bounding boxes to highlight regions of interest with minimal background noise, which suggests that such a model has immediate utility as a diagnostic supplement. In resource-restricted areas where access to radiologists may be limited, such a model could help patients by offering a diagnosis that can be easily reviewed by other available front-line workers. Additionally, our model can reduce the burden of overwhelmed, fatigued radiologists.

Our results demonstrate a diagnostic system that is more sensitive than RT-PCR testing, which previous studies have shown to have false negative rates as high as 30%.^6–8^ While previous AI studies for COVID-19 diagnosis have demonstrated effective preliminary results, model interpretability has been largely ignored, limiting clinical translation. Attempts at making models more transparent have focused on using activation maps but leave ambiguity as to how a diagnosis was assigned and lack the clarity needed for human verification.^15–18^ By focusing on interpretability, our model was also able to refine the previously suspected heterogeneity of GGOs. While our system has proven to be highly promising across all performance metrics, we are limited by the relatively small size of our dataset. More data is required to further refine this model and validate its robustness. To address this restriction, the model architecture can be easily adapted for future training through transfer learning, allowing for a rapid incorporation of more data. Further, our study is limited in basing diagnoses solely from radiological findings from chest CT scans. A recent report showed findings of uncharacteristic COVID-19 positive patients whose CT scans showed no GGOs.^14^ While these findings were limited, such patients may be inaccurately classified by our model. Finally, while the pathology of DQ features has been generally accepted,^12^ there may be highly irregular cases of COVID-19 that present with such phenotypes. In these events, the decision tree logic would incorrectly assign the patient as COVID-19 negative, although such cases are likely to be isolated. In summary, we presented a highly robust and interpretable model capable of diagnosing COVID-19. We also reported findings which indicate asymptomatic diagnosis is immediately possible and that GGOs are largely heterogeneous and disease-specific. With further study, CT scans of GGOs augmented by AI could potentially serve as an effective tool by which various diseases can be effectively diagnosed. Future studies may also find AI to be useful in asymptomatic diagnosis of not only COVID-19 but also of many other diseases. Combining CT scans with other clinically relevant material from a patient’s electronic health record may improve AI diagnosis of COVID-19. Further work is needed to normalize artificial intelligence in healthcare systems, and models such as this provide effective avenues to create a new diagnostic paradigm.

## Data Availability

Data and materials will be fully available upon reasonable request. Model weights and code will be made public.

## Acknowledgements

We would like to thank Dr. Gitanjali Vidyarthi for her thoughtful comments and suggestions as well as for providing introductions between authors. We would like to thank Zichen Miao for support on testing some visualization tools.

## Funding

S.S.M. is partially supported by a Veteran Affairs Research Career Scientist Award (IK6 BX003778), VA COVID Rapid Response Support (BX003685 Suppl), and University of South Florida Strategic Investment Program Fund. S.M. is partially supported by Research Career Scientist Award (IK6 BX004212). G.S. has research activities partially supported by the National Science Foundation (NSF 1712867), the Department of Defense (ONR N00014–18–1–2143-P00001, ONR N00014–20–1–233, NGA HM04761912010), the National Institutes of Health (1R01-MH122370–01, 1R01-MH120093–01), the Simons Foundation, and gifts from Microsoft, AWS, and Google.

## Author Contributions

A.W. and P.W. contributed equally to this work. A.W. and P.W. conceived of the study and A.W., P.W, R.W. designed, generated, and validated the model. A.W., A.S., P.P, L.C. gathered data for the model. A.S., P.P, N.V. labeled the data and reviewed labels to establish consensus. A.W., P.W., R.W., G.S., S.S.M., S.M. analyzed model and results. A.W. and P.W. wrote the paper. All authors discussed the results and commented on the manuscript.

## Conflicts of Interest

G.S. is a consultant for Apple and Volvo and has received speaker fees from Janssen on topics not related to this manuscript. The contents of this report do not represent the views of the Department of Veterans Affairs or the United States Government. The funders had no role in study design, data collection, and analysis, decision to publish, or preparation of the manuscript. All other authors declare no competing interest.

## Footnotes

* While we often discuss “diagnosis,” the tool here developed can be used for screening and/or complementing other techniques, including radiologists’ examination; as such, the results in this work should be considered for a broad range of applications and deployments.

## Notes

### Author Declarations

No IRB necessary

